# “This is a quiz” Premise Input: A Key to Unlocking Higher Diagnostic Accuracy in Large Language Models

**DOI:** 10.1101/2024.09.20.24314101

**Authors:** Yusuke Asari, Ryo Kurokawa, Yuki Sonoda, Akifumi Hagiwara, Jun Kamohara, Takahiro Fukushima, Wataru Gonoi, Osamu Abe

**Author notes:** Correspondence to Ryo Kurokawa, MD, PhD, Department of Radiology, Graduate School of Medicine, The University of Tokyo, 7-3-1, Hongo, Bunkyo-ku, Tokyo, 113-8655, Japan., Email address.

## Abstract

**Purpose:** Large language models (LLMs) are neural network models trained on vast amounts of textual data, showing promising performance in various fields. In radiology, studies have demonstrated the strong performance of LLMs in diagnostic imaging quiz cases. However, the inherent differences of prior probabilities of a final diagnosis between clinical and quiz cases pose challenges for LLMs, as LLMs had not been informed about the quiz nature in previous literature, while human physicians can optimize the diagnosis, consciously or unconsciously, depending on the situation. The present study aimed to test the hypothesis that notifying LLMs about the quiz nature might improve diagnostic accuracy.

**Methods:** One-hundred-and-fifty consecutive cases from the “Case of the Week” radiological diagnostic quiz case series on the American Journal of Neuroradiology website were analyzed. GPT-4o and Claude 3.5 Sonnet were used to generate top three differential diagnoses based on the textual clinical history and figure legends. The prompts included or excluded information about the quiz nature for both models. Two radiologists evaluated the accuracy of the diagnoses. McNemar’s test assessed differences in correct response rates.

**Results:** Informing the quiz nature improved the diagnostic performance of both models. Specifically, Claude 3.5 Sonnet’s primary diagnosis and GPT-4o’s top 3 differential diagnoses significantly improved when the quiz nature was informed.

**Conclusion:** Informing the quiz nature of cases significantly enhances LLMs’ diagnostic performances. This insight into LLMs’ capabilities could inform future research and applications, highlighting the importance of context in optimizing LLM-based diagnostics.

## Introduction

Large language models (LLMs) are neural network models trained on massive amounts of textual data, demonstrating excellent performance in various natural language processing tasks, including those in the medical field [1–3]. Recently, there have been attempts to apply LLMs to radiological diagnostics. Numerous studies have reported the use of LLMs in quiz cases for diagnostic radiologists. Reports indicate that by inputting textual data such as clinical history and image findings, ChatGPT’s GPT-4 model (OpenAI, San Francisco, United States) performed well in diagnostic imaging quizzes from radiology journals [4, 5]. Other studies have shown LLMs achieving good results in Radiology Board Examinations across different countries [6–8]. Comparative studies among vendors have also been conducted, with Sonoda and Kurokawa et al. [9] reporting that in text-based “Diagnosis Please” assessments, Claude 3 Opus (Anthropic, California, United States), GPT-4o (OpenAI, San Francisco, United States), and Gemini 1.5 Pro (Google, Mountain View, United States) scored significantly higher in this respective order. Efforts to use vision-language models that directly input radiological images are underway, and studies have reported improved accuracy in LLMs through key image input [10, 11].

However, when compared to human radiologists, Horiuchi et al.[12] reported that LLMs still perform lower than human radiologists in challenging neuroradiology cases. One notable issue is that the prior probability of a final diagnosis differs between clinical cases and quiz cases. Human radiologists, when presented with a quiz, tend to provide answers focusing on rare diseases, recognizing that these are more likely to be the intended answers. Conversely, they are inclined to avoid suggesting common diseases (such as cerebral infarction and subarachnoid hemorrhage) as these are less likely to be the quiz’s target. In contrast, the aforementioned studies did not inform the LLMs that they were dealing with quizzes, thus putting the LLMs at a disadvantage compared to humans.

We hypothesized that notifying LLMs that they are dealing with quiz cases for diagnostic radiologists might improve their diagnostic accuracy. The present study aimed to assess the impact of revealing the quiz nature of cases on the diagnostic performance of LLMs.

## Material and Methods

Ethical approval was not required as this study exclusively used publicly available information on the AJNR website (https://www.ajnr.org/cow/by/year).

We utilized the clinical history and figure legends from the “Case of the Week” series, a weekly quiz case compilation for diagnostic radiologists available on the American Journal of Neuroradiology website (https://www.ajnr.org/cow/by/year). A total of 150 consecutive cases from August 2021 to June 2024 were analyzed.

We used GPT-4o (OpenAI, San Francisco, United States; released on May 13, 2024) and Claude 3.5 Sonnet (Anthropic, California, United States; released on June 27, 2024) to list the primary diagnoses and two differential diagnoses for the cases.

Application programming interfaces were used to access each model (GPT-4o: gpt-4o-2024-05-13; Claude 3.5 Sonnet: claude-3-5-sonnet-20240620) on August 8, 2024. To ensure reproducibility, we specified the generation parameters for all models as temperature □ = □ 0.0. To prevent previous inputs from influencing subsequent ones, inputs were conducted in an independent session for each case for each condition. The prompt was as follows: “Assuming you are a physician, please respond with the most likely diagnosis and the next two most likely differential diagnoses based on the attached information” with or without the following sentences in the prompt to reveal the premise that they are quiz cases for experienced diagnostic radiologists (“additional prompt”) for LLM “They are quizzes of diagnostic imaging for doctors specializing in diagnostic radiology, and the purpose of the questions is to share knowledge on rare diseases and their imaging findings. Please keep this premise in mind and answer the questions, considering that common diseases are less likely to be asked.” Each prompt was submitted to the models only once, and the first response generated was used for evaluation.

The accuracy of the primary diagnosis and two differential diagnoses generated by the models were determined by consensus between one trainee radiologist and one board-certified diagnostic radiologist with 11 years of experience.

McNemar’s test was used to assess the difference in correct response rates for the overall accuracy under Condition 1 (without additional prompt) and 2 (with additional prompt) for each model and between the models. Two-sided p-values □ < □0.05 were considered statistically significant. Statistical analyses were performed using R (version 4.1.1; R Foundation for Statistical Computing, Vienna, Austria).

## Results

As shown in Table 1, Claude 3.5 Sonnet exhibited significantly superior diagnostic performance compared to GPT-4o under all conditions (p < 0.0001, McNemar’s test). Revealing the quiz context significantly improved the top three differential diagnoses for GPT-4o and the primary diagnosis for Claude 3.5 Sonnet (p = 0.020 and p = 0.0075, respectively). While improvements were observed in GPT-4o’s primary diagnosis and Claude 3.5 Sonnet’s top three differential diagnoses upon revealing the quiz context, these did not reach statistical significance.

**Table 1.**
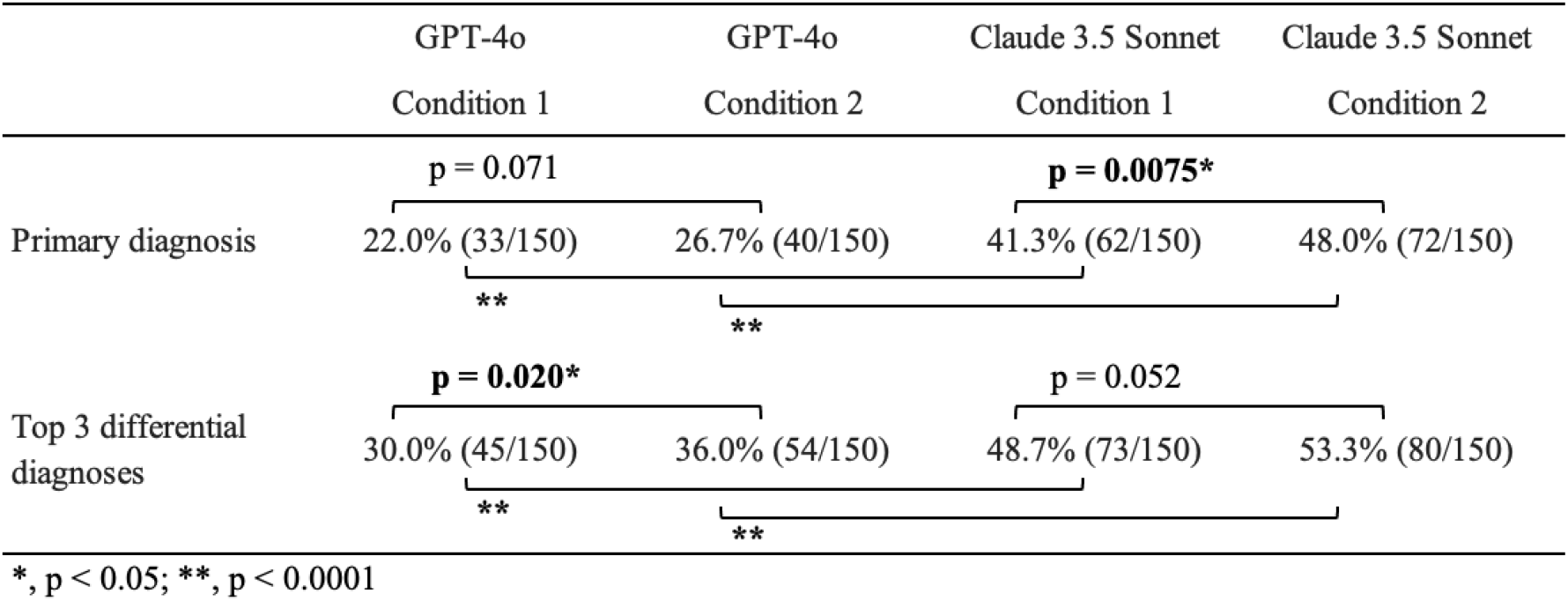
Diagnostic performances of large language models. All P-values shown are the results of McNemar’s test. (*, p < 0.05; **, p < 0.0001)

Specifically, Claude 3.5 Sonnet, without the quiz context, incorrectly proposed sinonasal lymphoma as the primary diagnosis for the February 9, 2023 case (https://www.ajnr.org/content/cow/02092023) but correctly diagnosed the rare Rosai-Dorfman disease when informed of the quiz nature. Similarly, GPT-4o failed to include the rare L-2-hydroxyglutaric aciduria in its differential diagnoses for the October 20, 2022 case (https://www.ajnr.org/content/cow/10202022) without information of the quiz nature, but correctly identified it as the primary diagnosis when informed of the quiz nature.

## Discussion

This study compared the diagnostic capabilities of different LLMs and explored the impact of informing them of the quiz nature of cases. Claude 3.5 Sonnet significantly outperformed GPT-4o in all conditions, and both models showed enhanced performance when aware of the quiz context.

Research involving LLMs solving quiz cases with definitive diagnoses is crucial for performance assessment of LLMs (inter-vendor comparisons, intra-vendor version comparisons), evaluating similarities and differences with human radiologists, and exploring future applications [9, 11]. However, existing studies did not inform LLMs of the quiz nature.

According to Bayes’ theorem, the pre-test probability (disease prevalence) is a critical determinant of post-test probability and subsequent diagnosis [13, 14]. Physicians, including radiologists, understand that disease frequency varies depending on the situation [15, 16].

Based on this premise—whether it’s a quiz case, regional differences, or the setting of a clinic versus third referral hospitals—they apply a gradient from high pre-test probability diagnoses to extremely low ones. Our assumption was that the same principle could apply to LLMs. We demonstrated that including the quiz context in the prompt significantly improved diagnostic performance.

Our study suggests future research directions: similar to how the quiz context enhanced diagnostic performance, presenting the clinical context might improve real-world diagnostic capabilities. It is known that appropriate clinical information enhances radiologists’ diagnostic accuracy [17]. Providing LLMs with clinical information databases (age and gender distribution, region, facility size, disease prevalence, etc.) could yield optimized diagnostic results for individual patients.

This study has some limitations. We did not analyze disease- or category-specific performance due to the limited number of cases. Additionally, the history and legends used are publicly available, possibly included in the training data of GPT-4o and Claude 3.5 Sonnet.

In conclusion, by informing the LLM that the case is a quiz case, the diagnostic performance based on text data from the history and figure legend significantly improved. Understanding this feature of LLMs could be valuable for future research and clinical applications.

## Data Availability

All data is available on the AJNR website (https://www.ajnr.org/cow/by/year).

